# A Meta-analysis of the efficacy of topical antibiotics in spinal surgery for the prevention of surgical site infection

**DOI:** 10.1101/2024.04.18.24305937

**Authors:** Yanfei Wang, Ke Song, Songlin Cai, Weifei Wu

**Author notes:** Corresponding author: Weifei Wu Corresponding Author’s Institution: ^1^the First College of Clinical Medical Science, China Three Gorges University;, ^2^Yichang Central People’s Hospital.

## Abstract

**Background:** Despite significant advancements in clinical aseptic techniques and wound infection control, surgical site infections (SSIs) continue to pose a significant risk and complication following spinal surgery. The use of intrawound antibiotics for the prevention of SSIs after spine surgery is a controversial method.

**Objective:** To conduct a review of the current literature on the use of antibiotics in wound care and evaluate their effectiveness in preventing postoperative SSIs.

**Methods:** Keywords such as “spinal surgery” or “spine”, “antibiotics”, “local” or “topical”, “prevention of infection”, and “infection” were used based on PubMed, Web of Science, Cochrane and Embase database. The literature was screened based on the title, abstract, full text reading, and extraction of relevant research data. Comparisons of the data were performed using RevMan 5.3 software.

**Results:** A total of 18922 patients from 24 studies were included in the final analysis, 8878 patients received antibiotics (experimental group) to prevent SSIs, and 10044 patients did not receive any additional antibiotics (control group). In the experimental group, 178 patients developed SSIs, compared to 356 patients in the control group. The results of the meta-analysis indicated that the incidence of SSIs in the experimental group was significant lower than that in the control group (95% confidence interval, 0.36-0.75, p=0.0004).

**Conclusion:** The topical application of antibiotics within the wound site is a crucial and efficient method to prevent SSIs after spinal surgery.

## 1. Introduction

Spinal surgical site infections (SSIs) are a significant and common complication that can result in severe health issues. The occurrence of SSIs after spinal surgery varies from 0.7 to 16.1%.^[1-2]^ Infections in spinal surgery can cause pain, fever, spinal deformity, neurological dysfunction, and even systemic infection, posing a grave risk to patients’ quality of life and survival rate.^[3]^ The severity of the infection depends on several factors, such as the type and location of the infection, the patient’s immune status, and the response to treatment.^[4-5]^ The removal of implants and subsequent surgery may be necessary in patients who have developed SSIs following spinal surgery, particularly in cases of chronic infection.^[6]^ Patients afflicting with such infections face significant costs in their treatment.^[7-8]^ Furthermore, in certain instances, death can also occur. There were 500,000 patients with SSIs in the United States each year, and the cost of treatment can be as high as $1.8 billion.^[9]^ Rational and effective use of antibiotics at the site of spinal surgery maybe significantly lower patient costs. According to Emonare, a study found that 207 patients who did not receive antibiotics within the surgical site incurred an additional cost of $573,897.92. The total cost of treatment for the 150 patients who received antibiotics inside the wound was approximately $1,152.^[10-11]^ Therefore, it is crucial to possess a comprehensive understanding of the seriousness of spinal SSIs and to implement efficient preventative and therapeutic measures.

There is still some controversy regarding whether the use of antibiotics at the site of spinal surgery has an effect. Some studies found the prophylactic use of antibiotics can significantly reduce infection rates or decrease the likelihood of staph infection after spinal surgery.^[12-19]^ However, other findings indicated that excessive use of antibiotics can result in an escalation of bacterial resistance, which could intensify the treatment challenges.^[20-21]^ Some studies suggested that there was no significant difference between using antibiotics and not using antibiotics in the outcome of an infection. When infection rates were low, antibiotics may not be effective.^[22-25]^ One study demonstrated that the use of antibiotics increased the rate of infection with gram-negative bacteria.^[25]^ Another research study indicated that antibiotics can reduce staphylococcal infections at surgical sites.^[26]^ In response to this controversy, we conducted a meta-analysis to summarize and analyze the existing literature on the effectiveness of antibiotics in preventing wound infection in spinal surgery, to help doctors make more informed decisions about use of antibiotics.

## 2. Methods

### 2.1 Literature retrieval

This meta-analysis was registered with PROSPERO (CRD:42024519225) and conducted according to PRISMA Guidelines. PubMed, Web of Science, Cochrane, and Embase databases were searched using keywords such as “spinal surgery” or “spine”, “antibiotics”, “local” or “topical”, “prevention of infection”, and “infection”. Randomized controlled trials (RCTs), prospective and retrospective studies, case reports, and observational studies were included in final analysis. This meta-analysis did not require approval from an Ethics committee. A total of 632 articles were retrieved from four retrieval repositories on the topics of removal conferences, meat analysis, animal experiments, and non-spinal surgery. Finally, 24 articles were selected for the study. The title, abstract author, publication journal, publication year, and other relevant information from the literature were extracted and recorded for further analysis.

### 2.2 Data extraction

Two researchers independently extracted bibliometric indicators and discussed the differences until a consensus was reached. Microsoft Excel was used to extract and analyze the data, including author, journal, year, title, study type, intervention method and sample size. The total number of patients, the number of patients receiving topical antibiotics, and the number of patients who developed SSIs were recorded as outcome parameters.

### 2.3 Analysis

The data were carefully reviewed to ensure accuracy. The extracted data and outcome measures of the study were analyzed and processed using RevMan 5.3 software. The study exhibited significant heterogeneity when I^2^> 50%. Consequently, the random effects of the research data were analyzed based on this situation. When I^2^< 50%, a fixed effects model was used to analyze the study data. The data from each study was analyzed using random effects analysis, and the differences in each study were defined using odds ratio (OR) values and a 95% CI to classify the experimental and control groups. The OR of all experiments was represented by a forest plot.

## 3. Results

### 3.1 Study selection

Out of a total of 632 studies, 24 studies comparing the effectiveness of topical use of antibiotics to prevent SSIs after spinal surgery were included in the final meta-analysis, 7 were randomized controlled trails, 17 studies were prospective and retrospective studies. The studies reported final outcomes of 18922 patients who underwent spinal surgery, either with or without antibiotics. Among these patients, 8878 received antibiotics for SSIs prophylaxis, while 10044 did not receive additional antibiotics (Figure 1 and Table 1). The evidence level of the studies ranged from II to III, and the NOS score ranged from 5 to 7 (Table 1).

**Table 1.** Demographic characteristics included in the study. RCT, randomized controlled trial; NIA, not available.

| Study design | Author | Year | antibiotics/control | average ages<br>(years) | type of surgery | dose<br>(g) | follow-up<br>(month) | Evidence | Quality |
| --- | --- | --- | --- | --- | --- | --- | --- | --- | --- |
| Prospective | Sohrab Salimi | 2022 | 187/188 | 51.7/52.4 | spinal surgery | 1-2 | 3 | III | 7 |
| Prospective | Clinton J. Devin | 2015 | 966/1090 | 60.5/59.5 | NA | 1 | NA | II | 7 |
| Prospective | Leslie | 2008 | 116/117 | NA | lumbar fusion | 1-2 | NA | III | 6 |
| RCT | Guilherme Finger | 2020 | 49/47 | 43 ± 14.88 | spinal fusion | 2 | 5-24 | III | 7 |
| RCT | Shah Khalid | 2023 | 39/39 | 36.9/39 | spinal surgery | 1 | 3 | III | 7 |
| RCT | B. Mirzashahi | 2018 | 193/187 | NA | NA | 1-2 | 15 | II | 6 |
| RCT | Vijay Ramappa<br>Tubaki | 2013 | 433/474 | 44.5/46.6 | NA | 1 | 3 | II | 7 |
| RCT | Ralph T. Schär | 2021 | 17/17 | 63.5/60.5 | spinal fusion | 2 | 1.5 | III | 6 |
| RCT | Mikinobu<br>Takeuchi | 2018 | 230/0 | 67 | horacic or lumbar<br>fusion | 1 | 12 | III | 6 |
| RCT | Petignat | 2008 | 616/624 | 46/45 | herniated disc | NA | 3-6 | II | 7 |
| Retrospective | Krishn Khanna | 2019 | 2357/2521 | 61.2/58.1 | spinal surgery | 1-2 | NA | III | 6 |
| Retrospective | Marian L. Gaviola | 2016 | 116/210 | 62/55 | spinal fusion | 2 | 3 | III | 7 |

Table 1. Continued.
| Study design | Author | Year | antibiotics/control | average ages<br>(years) | type of surgery | dose<br>(g) | follow-up<br>(month) | Evidence | Quality |
| --- | --- | --- | --- | --- | --- | --- | --- | --- | --- |
| Retrospective | Aaron Heller | 2015 | 342/341 | 55.3/49.1 | NA | 0.5-1 | 3 | III | 7 |
| Retrospective | Hey Hwee Weng<br>Dennis | 2017 | 117/172 | 45/48 | NA | 1 | 3 | III | 6 |
| Retrospective | Hill | 2013 | 150/150 | 54.1/58.3 | NA | 1-2 | 1 | III | 5 |
| Retrospective | Marko Tomov | 2015 | 1173/1252 | 57.4 | NA | 1 | NA | III | 6 |
| Retrospective | Osa Emohare | 2014 | 96/207 | 53.7/58.2 | NA | 1 | NA | III | 7 |
| Retrospective | Shoichi Haimoto | 2018 | 247/268 | 58.4/54.4 | NA | 1 | NA | III | 7 |
| Retrospective | Tetsuro Hida | 2017 | 81/93 | 48.4/50.3 | NA | 0.5-1 | 20 | III | 6 |
| Retrospective | Sweet | 2011 | 911/821 | 56/53 | Lumbar/thoracic | 2 | 30 | III | 7 |
| Retrospective | Godil | 2013 | 56/54 | 43/45 | Fracture | 1 | 3 | III | 6 |
| Retrospective | Joel R. Martin | 2015 | 115/174 | 62.3/57.6 | Cervical | 2 | NA | III | 6 |
| Retrospective | Pahys | 2011 | 195/806 | 57.1/54.9 | cervical | 0.5 | NA | III | 6 |

**Fig. 1.**
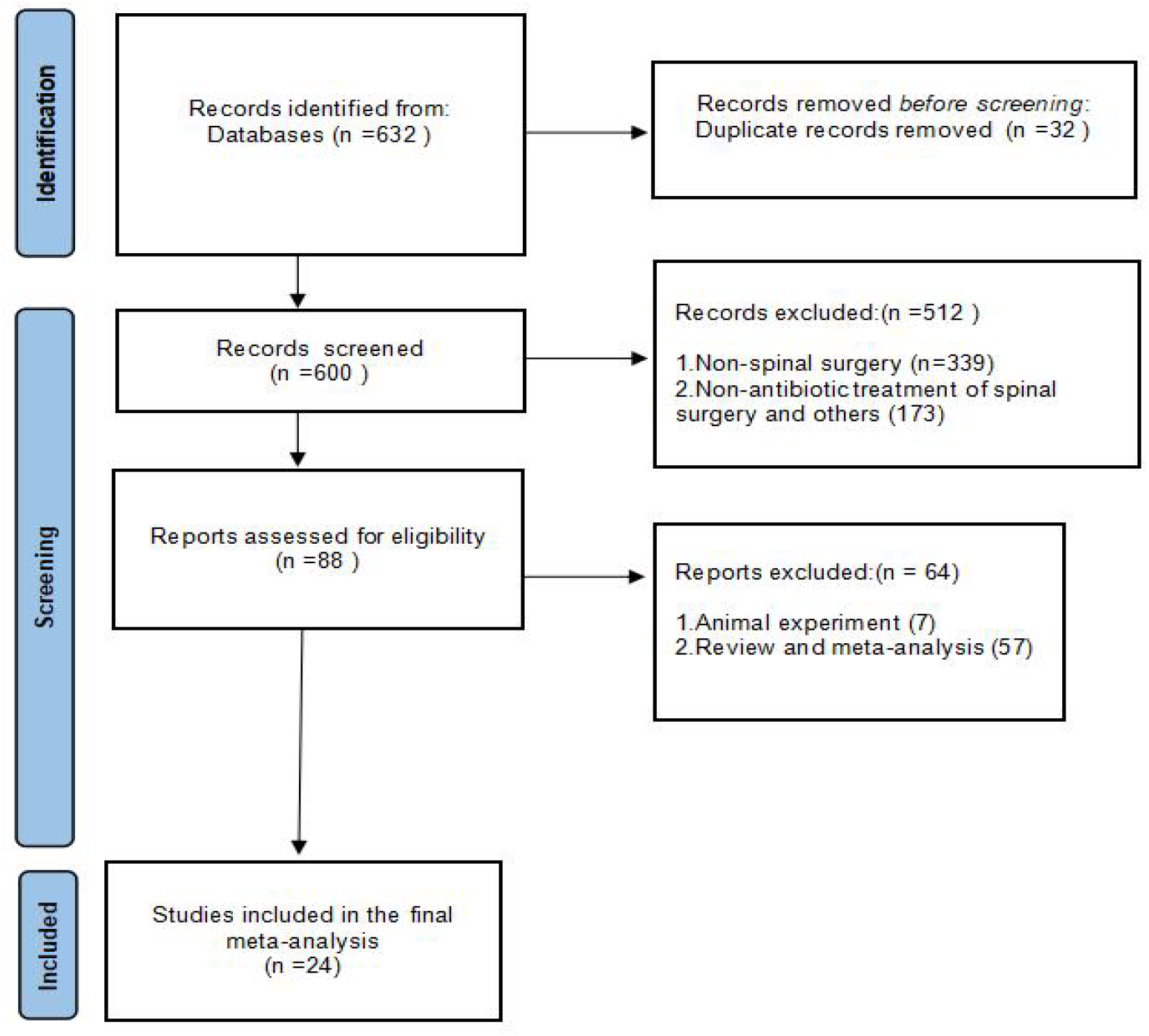
A total of 24 research articles were selected for inclusion in the final study.

### 3.2 Over analysis

The effectiveness of antibiotics in preventing SSIs after spinal surgery was found to be 59% (I^2^ = 59%). The overall OR was 0.52 (95% confidence interval [CI]=0.36-0.75), indicating that the incidence of SSIs in the antibiotic treatment group was significantly lower than that in the control group (p=0.0004, Figure 2). Additionally, we included a study that did not have a control group but compared the efficacy of two different antibiotics.^[27]^

**Fig. 2.**
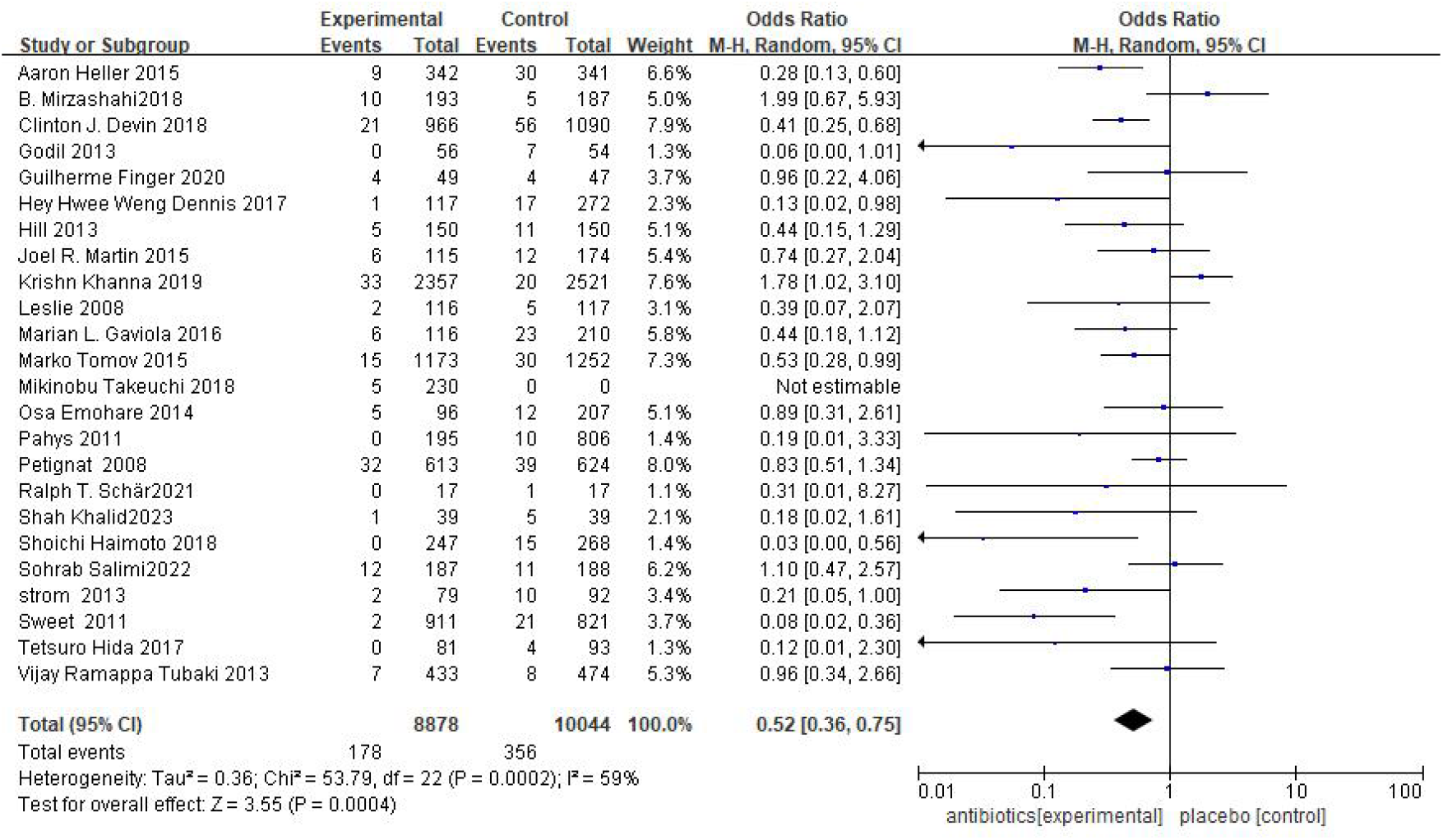
The infection rate of surgical site in the local antibiotic treatment group was significantly lower than that in the control group.

### 3.3 Subgroup analysis according to the study design

Compared to randomized controlled trials, retrospective studies are susceptible to information bias and confounding factors during data analysis, which may introduce a certain degree of bias to the results. In this study, we analyzed the results of randomized controlled trials, retrospective studies, and other types of studies. Specifically, 7 randomized controlled trials were included, which involved a total of 3399 patients (1782 patients receiving antibiotics and 1617 patients in the control group). Overall, the OR=0.75 with a 95% CI=0.52-1.07 (Figure 3). The results of the meta-analysis indicated that the incidence of SSIs in the antibiotic group was 0.75 times higher than that in the non-antibiotic group. However, there was no significant difference between the two groups (p=0.13). Additionally, the meta-analysis of non-randomized controlled trials (non-RCTs) studies showed that the incidence of SSIs in the antibiotic group was 0.43 times higher than that in the control group. Furthermore, data from 17 prospective, retrospective, and case studies involving 15960 patients (7304 in the antibiotic group and 8656 in the control group) demonstrated that SSIs in the antibiotic group was obvious lower than SSIs in the control group (OR=0.43, 95% CI=0.27-0.66, p=0.0002, Figure 4).

**Fig. 3.**
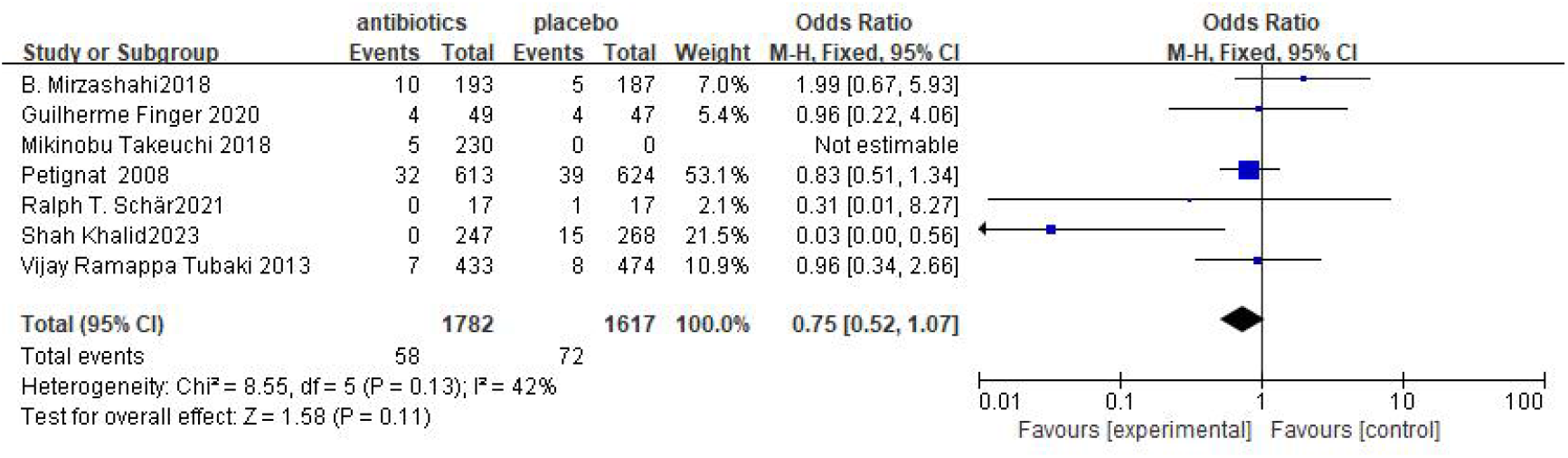
Meta-analysis of randomized controlled trials (RCTs).

**Fig. 4.**
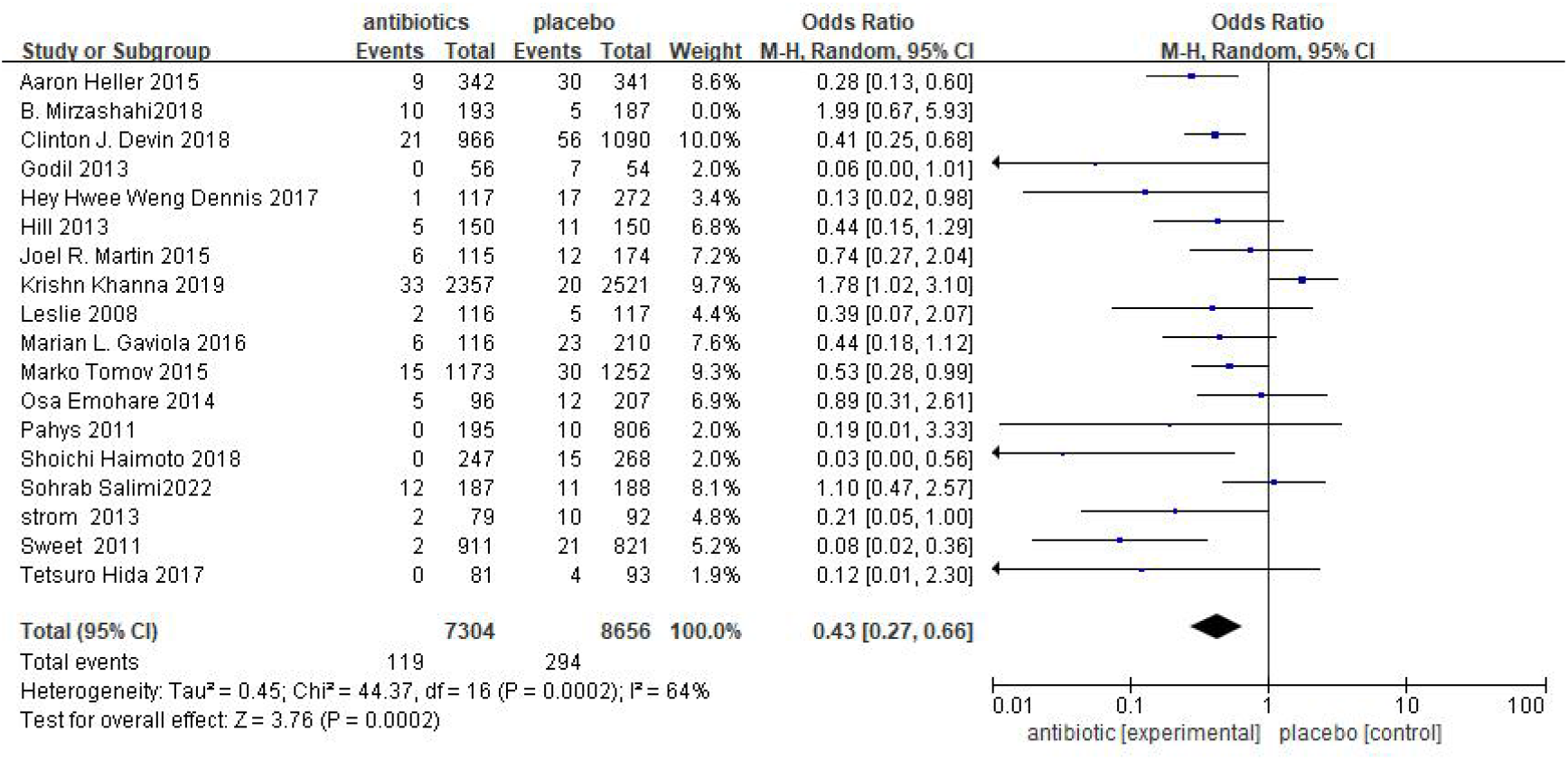
Meta-analysis results of retrospective and other studies.

### 3.4 Subgroup analysis according to the application of implants

Spinal fixation surgery is an important method for treating spinal diseases. However, complications such as infection may occur after surgery due to the use of foreign implants in the human body. This study analyzed 10 internal fixation studies involving a total of 5961 patients, with 2606 patients receiving antibiotics and 3355 control patients. The group receiving antibiotics had a lower incidence of SSIs compared to the control group. The incidence of SSIs was 0.26 times higher in the antibiotic group than in the non-antibiotic group, and this difference was statistically significant (OR=0.26, 95% CI=0.12-0.55, P=0.0004, Figure 5).

**Fig. 5.**
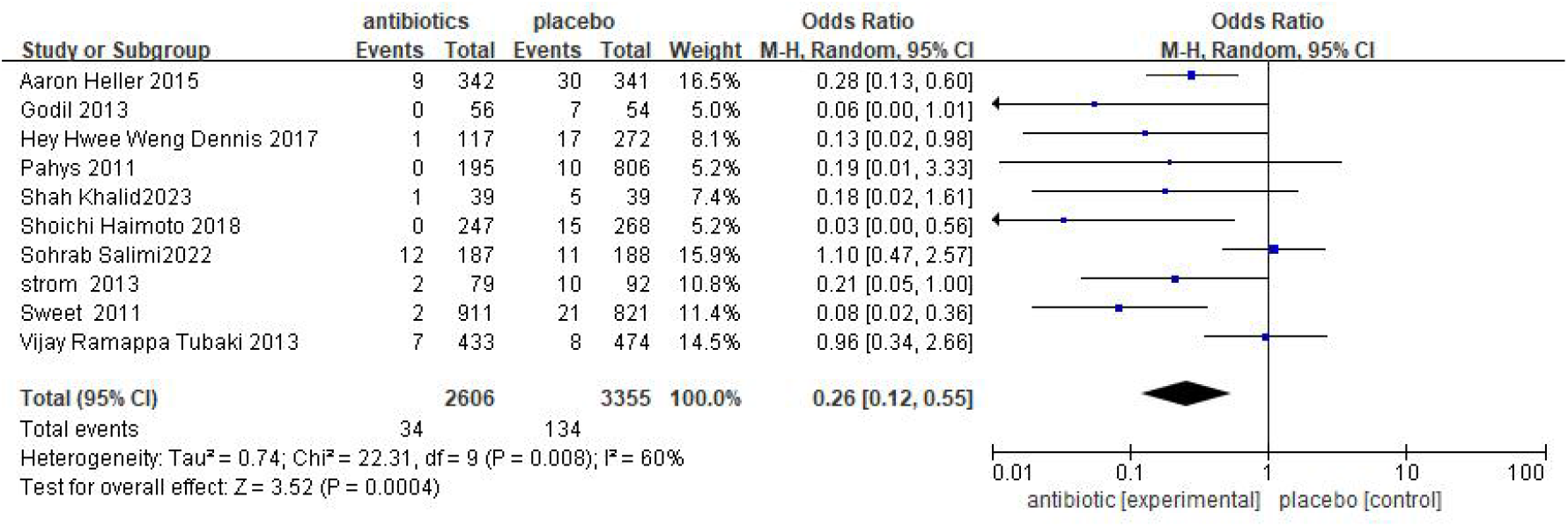
Results of meta-analysis using implants surgery.

### 3.5 Subgroup analysis of multiple microbial infections and Gram-negative bacteria

In some of the studies included, it is possible to obtain information on the incidence and severity of different bacterial infections after antibiotic use. Additionally, it is possible to compare the inhibitory effect of different antibiotics on various bacterial communities and evaluate the impact of antibiotic on wound infection. In this analysis, five studies were found that reported multiple microbial infections, which included a total of 6560 patients, with 3239 receiving antibiotics and 3321 serving as controls. The results of the meta-analysis showed OR=0.46, with a 95%CI=0.16-1.37 (Figure 6). The analysis indicated that the incidence of multiple organisms in the antibiotic group was 1.66 times higher than that in the control group. These results suggested that the use of antibiotics was not effective in reducing the infection of multiple microorganisms. There was no significant difference between the two groups. The results of the Gram-negative bacteria infection showed OR=1.58 with 95% CI=0.76-3.28 (Figure 7). The incidence of Gram-negative bacilli infection in the antibiotic group was 0.46 times higher than that in the control group. This indicates that patients who did not receive antibiotics were more likely to be infected with Gram-negative bacteria.

**Fig. 6.**
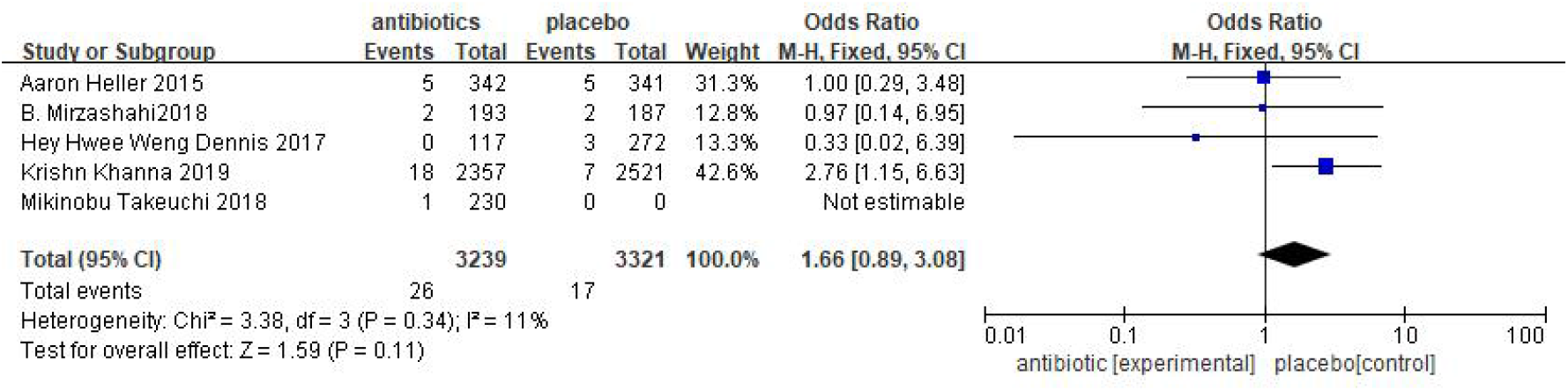
Results of meta-analysis of infection with multiple microorganisms.

**Fig. 7.**
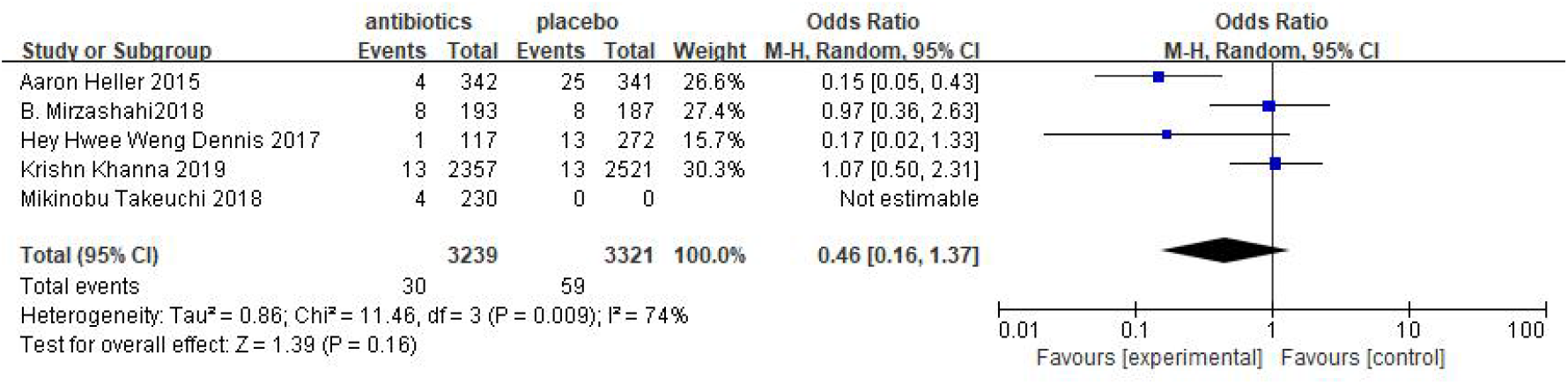
Results of meta-analysis of Gram-negative bacteria infection.

## 4. Discussion

Spinal surgery is a common surgical procedure, and SSIs are a serious complication that can increase the pain and cost of treatment for patients.^[28]^ Despite substantial advancements in clinical practice to prevent infections in spinal surgery, the risk of SSIs remains in clinical work. In this study, we investigated the efficacy of using local antibiotics to prevent SSIs in spinal surgery. Our research findings demonstrated that the use of local antibiotics reduced the risk of SSIs in patients undergoing spinal surgery. This suggests that local antibiotics have a certain level of effectiveness in preventing SSIs, which can ultimately enhance surgical outcomes and promote patient recovery.

The majority of infections after spinal surgery happen in the early postoperative period, typically within three months. Preoperative risk factors that have shown statistical significance include age over 60 years, smoking, previous surgical infection, diabetes, obesity, and alcoholism.^[29]^ Perhaps the use of spinal surgery implants is one of the factors that can contribute to the occurrence of SSIs. Nowadays, the use of implants has become a common practice in spinal surgeries, for diseases like vertebral fractures, spinal injuries, and degenerative spinal conditions. Implants can help restore the normal structure and function of the spine, stabilize it, and prevent abnormal movements. They can also help correct spinal deformities, improve abnormal posture, and alleviate pain symptoms. Bone graft materials and fillers can promote bone healing and fusion, making the healing of the spinal area more secure. Artificial intervertebral discs can restore the function of damaged discs to maintain normal spinal movement. These benefits make implants an essential and integral part of these surgeries, but the risk of postoperative infections caused by implants should not be ignored. Due to the potential formation of bacterial biofilms, which can attach to implants and protect pathogens from the host’s immune system and systemic antibiotics, they can cause difficult-to-treat SSIs and hinder wound healing.^[30]^ Patients with malnutrition, low resistance, weakened immune systems, or other diseases may have an increased risk of SSIs when using implants. In the multivariate analysis, being female (OR=3.3, p<0.01) and having diabetes (OR=0.51, p<0.01) were identified as significant risk factors for SSIs.^[31-32]^

The current routine route of administration for antibiotics is still intravenous systemic administration.^[33-34]^ There are several disadvantages to administering antibiotics through intravenous injection, including the reduction of antibiotic concentration in the targeted area, the inability to reach tissues with poor blood supply, and the potential for systemic toxicity,^[35]^ In contrast, local antibiotics can achieve high concentrations in local tissues while maintaining lower levels in the body, thereby avoiding potential side effects like kidney or ear toxicity.^[36]^ In the study conducted by Gupta S et al., it was concluded that the utilization of vancomycin in surgical incisions significantly decreased the occurrence of soft tissue infection (44.4% vs 100%) and implant infection (27.8% vs 100%). Furthermore, the use of topical antibiotics proved to be more effective than systemic intravenous antibiotics in preventing SSIs.^[37]^ In a case-control study conducted by Pinter Z et al., there were 316 cases of SSIs out of 19,081 spinal surgeries, resulting in an infection rate of 1.7%. The proportion of gram-negative bacilli was found to be 6%. Notably, the study revealed a significant decrease in SSIs among spine surgery patients who received intrawound vancomycin treatment.^[38]^

The topical antibiotics used after spinal surgery include vancomycin, cefazolin, cefuroxime, and ampicillin. The most commonly used is vancomycin powder, which is applied directly to the wound during the operation. Different antibiotics may have varying side effects in the treatment and prevention of SSIs. In a retrospective cohort study conducted by Tafish et al., 81 patients were administered vancomycin powder for SSIs prophylaxis, while 375 patients did not receive vancomycin. The results indicated that there were 8 SSIs in the treatment group and 20 SSIs in the control group. Although it was concluded that vancomycin powder did not reduce the incidence of SSIs in spinal surgery, it is important to note that the treatment group had 72 implants and the control group had 184 implants. Additionally, 71 patients in the treatment group and 29 patients in the control group had prolonged operation time. These factors are undeniably significant and may greatly influence the occurrence of SSIs.^[39]^ Therefore, selecting the appropriate antibiotics and dosage is crucial for preventing SSIs. In the study conducted by Xu S et al., a total of 192 cases of lumbar fusion were treated with vancomycin in order to prevent infection. The incidence of SSIs in the vancomycin group was 0.0%, which was significantly lower compared to the control group (5.3%). Additionally, no adverse events associated with the use of vancomycin powder were reported.^[40]^ A retrospective study conducted by Kadir Okta et al. examined the use of vancomycin powder in preventing SSIs during spinal instrument surgery in high-risk patients.^[41]^ Additionally, a study by Shiyong Wang et al. demonstrated the effectiveness of vancomycin powder in reducing the occurrence of SSIs at posterior depth during lumbar disc fusion.^[42]^ Furthermore, these two studies suggest that vancomycin could potentially prevent SSIs in patients who are older, have longer surgery durations, higher BMI, and diabetes.

As the latest studies have demonstrated that the use of vancomycin powder in the wound may increase the infection rate of gram-negative bacilli.^[25]^ In the study conducted by Hu W et al., it was observed that the bacterial flora of the infection changed after the occurrence of SSIs following the use of vancomycin powder at the spinal surgical site. The proportion of Gram-negative bacilli in the group infected with vancomycin powder was 46.4% in SSIs, whereas the proportion of Gram-negative bacilli in the group infected with non-vancomycin powder was 30.1%.^[43]^ According to the data of this analysis, the dosage of local antibiotics used in most studies was primarily controlled within the range of 1 to 2g. In cases where the skin incision exceeded 20cm, 2g of antibiotics were administered to prevent infection in the wound.^[16]^ Hyoda Y et al. demonstrated that the levels of C-reactive protein were significantly higher in the vancomycin group compared to the control group on postoperative days 1 and 3. They also found that the application of vancomycin powder topically resulted in an acute inflammatory response.^[43]^ Additionally, administering less than 1g of vancomycin powder topically during minimally invasive posterior lumbar disc fusion may decrease the likelihood of SSIs.^[44]^ Further studies are required to ascertain the ideal dosage of antibiotics following spinal surgery. In the study conducted by Gupta et al., they discovered that the incidence of SSIs varied among implants made from different materials. Specifically, cobalt-chromium alloy implants were found to be more susceptible to infection compared to stainless steel and titanium implants.^[37]^ In the future studies can further explore the application effects of various types of topical antibiotics in spinal surgery and compare the efficacy and safety. Additionally, further exploration is needed in the future to determine whether the relationship between spinal implants made of different materials and various types of antibiotics contributes to the occurrence of SSIs. These findings will aid in refining strategies to prevent SSIs during spinal surgery, ultimately enhancing surgical success and improving the quality of patient recovery.

## 5. Conclusions

Using topical antibiotics after spinal surgery is beneficial in preventing SSIs. Topical antibiotics can directly target the infection site, effectively decreasing bacterial growth and lowering the risk of infection. Additionally, they can also decrease the need for systemic antibiotics, reducing the chances of adverse reactions and antibiotic resistance. Consequently, employing topical antibiotics after spinal surgery is an effective approach for preventing SSIs when appropriate.

## Data Availability

All data generated or analyzed during this study are included in this article.

## Declarations

### Ethics approval and consent to participate

The review boards of the China Three Gorges University approved the present study.

### Consent for publication

The authors report no conflict of interest concerning the materials or methods used in this study or findings specified in this paper. The authors have reviewed the final version of the manuscript and approve it for publication

### Availability of data and material

All data generated or analyzed during this study are included in this published article

### Author Contributions

Conceptualization: WF W, Methodology: YF W, K S, SL C, Software: YF W, K S, Formal Analysis: YF W, Investigation: YF W and WF W, Resources: WF W, Original Draft Preparation: YF W, K S, SL C, Review & Editing: WF W, Supervision: WF W, Project Administration: YF W, Funding Acquisition: WF W.

## Acknowledgements

Not Applicable

